# A Deep Learning–Enabled Single-Cell Morpholomic Atlas of Nasal Swabs Distinguishes Chronic Inflammation from Sinonasal Malignancy

**DOI:** 10.64898/2026.01.09.26343551

**Authors:** Brittany T. Rupp, Andreja Jovic, Theresa Weaver, Kiran Saini, Meghan Burr, W. Jared Martin, Quinn T. Easter, Adam J. Kimple, Kevin Matthew Byrd

## Abstract

**Background:** Sinonasal malignancies frequently present with symptoms overlapping chronic inflammatory conditions such as chronic rhinosinusitis (CRS), complicating early detection and delaying treatment. A fast, scalable, non-invasive approach capable of resolving immune and epithelial cell states across inflammatory and malignant disease from routine nasal swabs could substantially improve clinical screening, leading to the initiation of appropriate treatment.

**Methods:** We developed a deep learning–enabled single-cell morpholomic framework using the REM-I platform to generate a reference atlas of >641K cell brightfield images from purified immune cell populations. This reference atlas was applied to >2.5 million images obtained from nasal swabs spanning a clinical spectrum of health, CRS, and sinonasal carcinoma. Embeddings were integrated using dimensionality reduction for differential feature testing and comparative feature enrichment across disease states.

**Findings:** Across the disease continuum, sinonasal carcinoma samples exhibited distinct immune remodeling, including increased myeloid-like cell abundance and elevated small dark pixel intensity consistent with enhanced granulocyte activity. Basophil/NK-enriched clusters contained tumor-associated cells with deep learning–derived morphologic signatures not observed in CRS or healthy samples. Tumor-associated epithelial cells were significantly smaller and displayed disease-specific morpholomic patterns distinct from chronic inflammation.

**Conclusions:** This study establishes a deep learning–enabled single-cell morpholomic atlas of nasal swabs spanning healthy epithelium, chronic inflammation and sinonasal malignancies. Morpholomic cytology reveals reproducible immune and epithelial states associated with inflammatory and malignant disease and provides a scalable, non-invasive framework for cellular stratification in sinonasal pathology, supporting future applications in early point-of-care diagnostics.

## INTRODUCTION

The practice of examining disease through individual cells has evolved alongside advances in microscopy, moving from early visualization of the cellular world described by Hooke, Schleiden and Schwann to clinically transformative approaches pioneered by Papanicolaou, and later extended through fine-needle aspiration techniques by Martin and Ellis that now enable minimally invasive diagnosis in routine clinical practice^1^. Modern cytopathology (or cytology) is an important medical tool used daily throughout clinics and hospitals to screen for and diagnose a variety of conditions, including infectious diseases, inflammatory disorders, and malignancies^2^. Samples can be collected using multiple techniques, including tissue biopsies, fine needle aspirations, blood draws, and tissue brushing; however, fast, routine, and reliable implementation of these techniques depends on procedural simplicity, patient comfort, diagnostic performance, and integration into established clinical workflows. These factors define both cytology’s promise and its practical limitations in contemporary clinical practice^3^.

Among current cytologic procedures, tissue brushing is among the least invasive approaches and can be performed in the clinic setting with minimal training. The most well-known example of this is cytologic brushing of the cervix (i.e., Papanicolaou tests or Pap smears)^4^, which has transformed care by allowing clinicians to identify abnormal cells early in the disease process. Despite this successes, comparable brush-based cytologic approaches have not been widely adopted across other anatomical sites or disease contexts, limiting the broader application of minimally invasive cellular diagnostics and underscoring the need for advanced, technology-enabled strategies to modernize cytology for contemporary clinical practice^5^.

Within the upper airway, the nasal cavity represents a setting where the limitations of contemporary cytology are particularly evident. Nasal cytology via tissue brushing has been primarily applied to investigate immune infiltration and epithelial remodeling in disease states including chronic rhinosinusitis (CRS)^6^, post-viral olfactory dysfunction^7^, and allergic rhinitis^8^, conditions in which inflammatory remodeling and shifts in eosinophil populations are well described. However, sinonasal malignancies present a distinct and unresolved diagnostic challenge because they are rare, diverse, lack routine screening strategies, and frequently present with symptoms and features that overlap with CRS and inflammatory nasal polyps^9^. Prolonged inflammation in CRS can further obscure early malignant changes, complicating timely diagnosis and contributing to the limited clinical adoption of nasal brushings for tumor detection^10^. Additionally, inflammatory nasal polyps mimic tumor-associated symptoms, increasing diagnostic ambiguity in routine clinical evaluation^11^. Methods including tissue biopsy and advanced imaging such as magnetic resonance imaging (MRI) and computed tomography (CT) are often necessary for definitive tumor diagnosis but are invasive, costly, and typically utilized too late, leading to two-thirds of sinonasal squamous cell carcinomas being diagnosed at an advanced stage (i.e. T3 or T4)^12–15^. Therefore, although the sinonasal cavity is readily accessible for minimally invasive sampling, new approaches must be explored to achieve the necessary resolution required to reliably distinguish chronic inflammation from malignancy.

Recent technological advances are reshaping cytology from a largely qualitative, labor-intensive practice into a scalable, quantitative discipline capable of supporting next-generation diagnostics^16^. Innovations in high-throughput imaging and microfluidics, have begun to address long-standing barriers to cytologic analysis, including inter-site staining variability, dependence on expert manual interpretation, and limited throughput^17^. Artificial intelligence and machine learning (AI/ML) approaches further enable automated extraction of both classical morphometric features and higher-order representations learned directly from large image datasets, capturing subtle cellular phenotypes that are difficult to resolve by conventional microscopy^18,19^. These advances provide a foundation for modernizing cytology as a scalable analytic framework capable of resolving disease-associated cellular states across diverse tissues and clinical contexts, including anatomically complex and diagnostically ambiguous sites such as the nasal cavity.

To explore if next-generation cytologic analysis can distinguish between sinonasal states (i.e. health, chronic inflammation, malignancy), we applied a deep learning–enabled single-cell morpholomic framework to nasal swab specimens spanning each state. Using the high-throughput, label-free brightfield imaging platform REM-I, we profiled more than 2.5 million individual cells obtained from minimally invasive nasal swabs collected from healthy controls, patients with CRS with polyps, and individuals with biopsy-confirmed sinonasal carcinoma. This data was contextualized using a reference atlas generated from over 641,000 images of annotated purified immune cell populations, providing lineage-informed grounding for downstream analyses. For each cell, we integrated human-interpretable morphometric features (including size, shape, texture, and pixel intensity–based measurements) with deep learning–derived features learned directly from raw cellular images, enabling unbiased, high-dimensional characterization of immune and epithelial phenotypes.

By integrating these features into a shared morpholomic space, we constructed a scalable single-cell atlas that captures cellular states across the continuum of health, chronic inflammation, and malignancy. Using this framework, we identify immune and epithelial morpholomic states associated with inflammatory remodeling and malignant transformation, revealing cellular features that distinguish carcinoma from chronic inflammatory disease despite overlapping clinical presentation. This work establishes a deep learning–enabled single-cell morpholomic atlas of nasal swabs and demonstrates the potential of morpholomic cytology as a non-invasive, scalable approach for cellular stratification in sinonasal pathology, with broader implications for modernizing cytologic analysis across diverse anatomical sites and disease contexts.

## RESULTS

### High-throughput imaging of nasal swabs reveals single-cell morpholomic features differences

We applied high-throughput, label-free single-cell morpholomic imaging to nasal swab specimens to profile immune and epithelial populations from routinely accessible clinical samples. Nasal swabs were obtained using a cytobrush (Medscand Cytobrush Plus) collected under an approved IRB protocol (#UNC-IRB-17-2677) from healthy controls (n=5), patients with chronic rhinosinusitis (CRS; n=5), and individuals with biopsy-confirmed sinonasal carcinoma (n=3) (Figure 1a; Table 1). Most CRS cases presented with nasal polyps, reflecting chronic inflammatory remodeling that clinically overlaps with malignancy. Each nostril was sampled independently to assess intra-patient variability and minimize cross-contamination.

**Figure 1:**
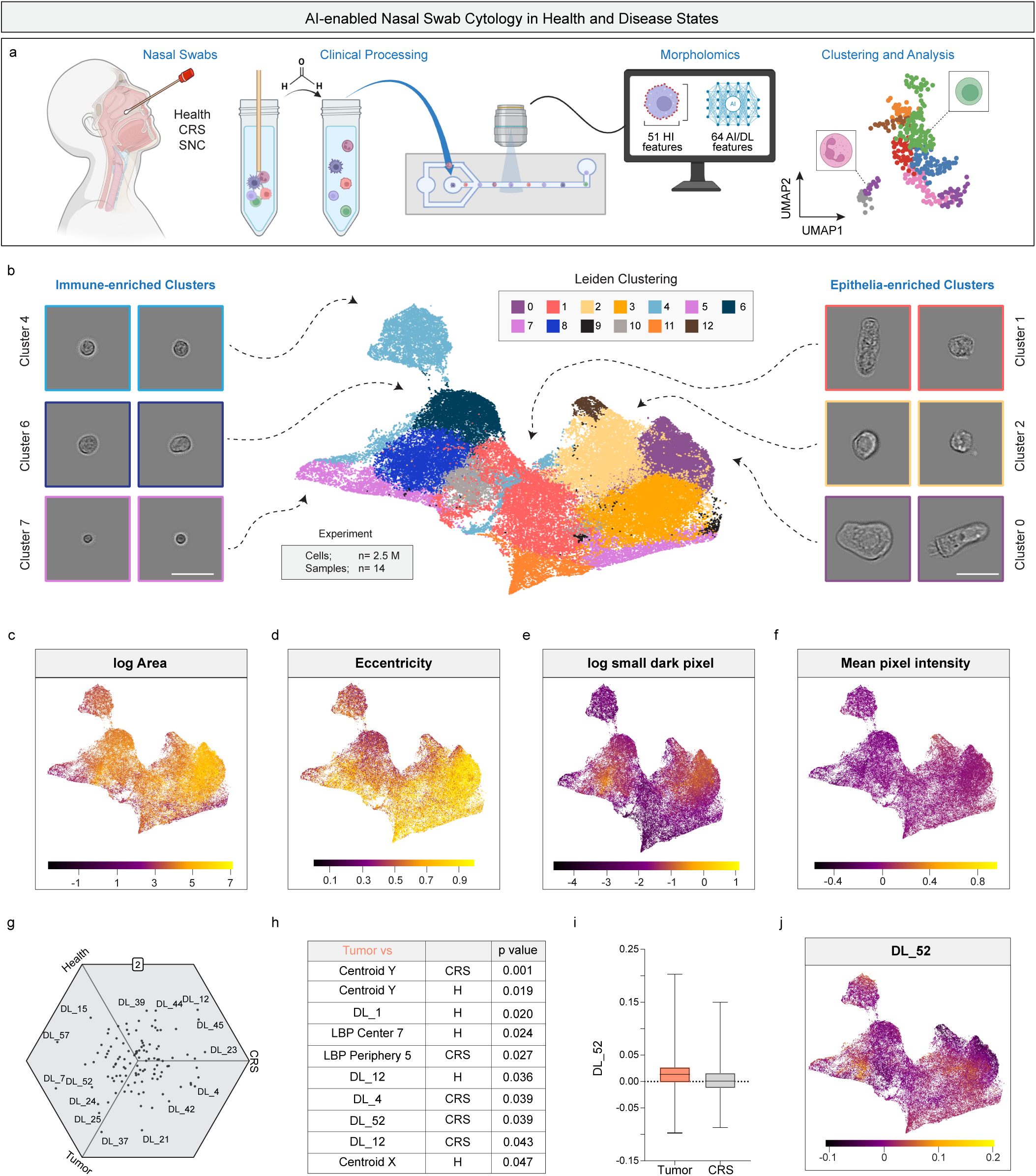
Overview of nasal swab morpholomic atlas with initial differential feature analysis. (a) Schematic of nasal swab collection, clinical processing, and high-throughput, label-free brightfield imaging followed by morpholomic feature extraction and unsupervised clustering. Chronic rhinosinusitis (CRS), Sinonasal Carcinoma (SNC) (b) UMAP embedding of nasal swab–derived cells (≈2.5 million cells from 14 samples) based on morpholomic features, with representative immune-enriched and epithelial-enriched cell images shown. Scale bar represents 20µm. (c–f) UMAPs colored by selected morpholomic features: (c) log-transformed cell area, (d) eccentricity, (e) log small dark pixel intensity (granule-associated texture feature), and (f) mean pixel intensity. (g) Tri-wise differential feature analysis comparing healthy, chronic rhinosinusitis (CRS), and sinonasal carcinoma samples using sample-level mean z-score–normalized features. (h) Results of Welch’s t-tests comparing median feature values in tumor samples versus CRS or healthy controls. (i) Distribution of DL_52 values in tumor and CRS samples. (j) UMAP colored by DL_52, illustrating spatial localization of tumor-associated deep learning features. Illustrations from a) were created with BioRender.com

**Table 1:**
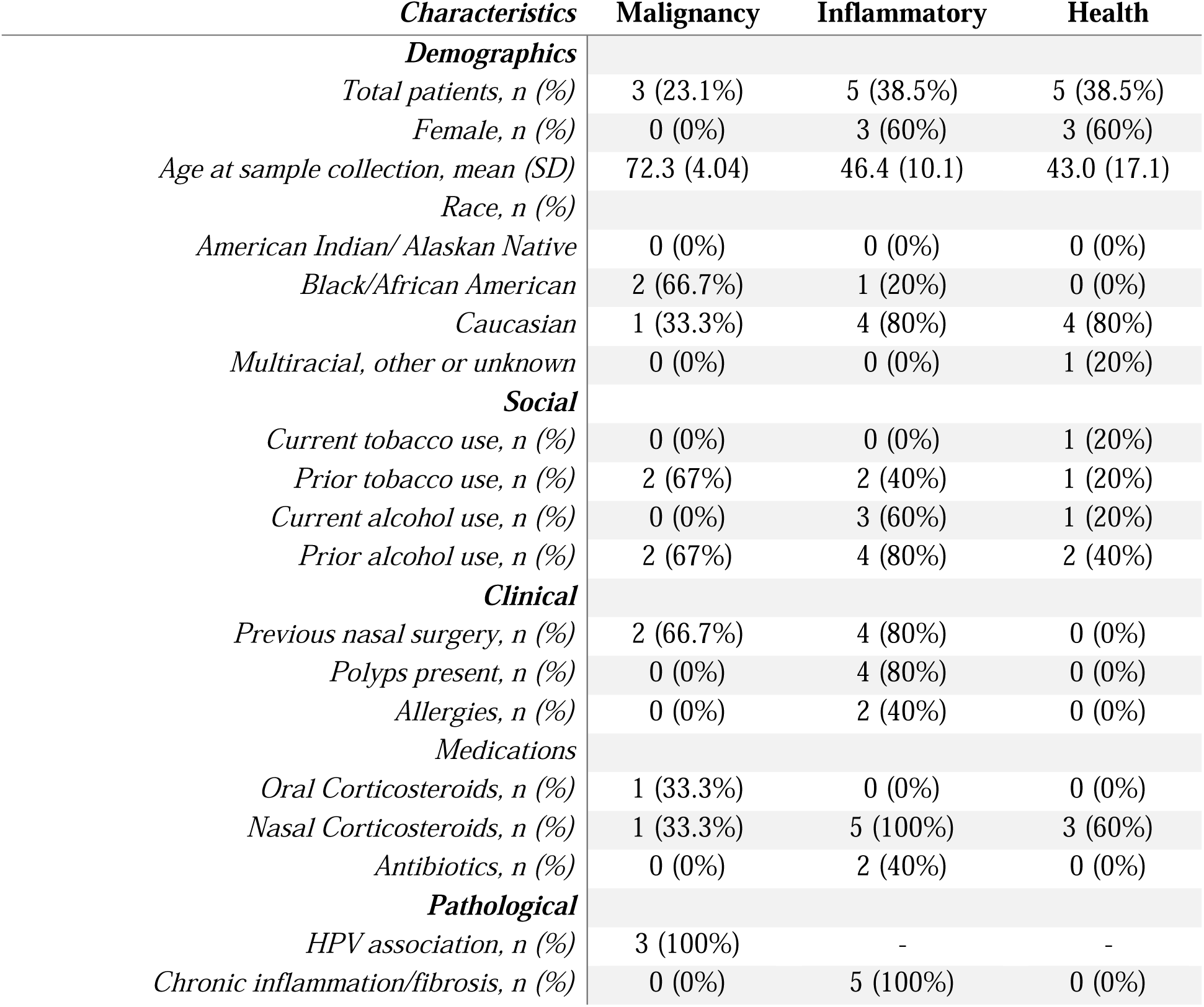
Demographic and clinical information.

Across all samples, more than 2.5 million individual cells were profiled using the REM-I platform, with 92.9% of swabs yielding at least 10,000 single-cell images (mean 90,998 images per swab, 690 cells imaged per minute). Real-time image processing extracted 115 morpholomic features per cell, including human-interpretable measurements of size, shape, intensity, and texture and deep learning (DL) derived features learned directly from cellular images. This workflow enabled scalable, unbiased morpholomic profiling across health, chronic inflammation, and carcinoma. Low-dimensional embedding of all nasal swab–derived cells revealed clusters with distinct morphologic characteristics consistent with immune and epithelial populations. While broad cellular classes could be inferred from size and shape, precise lineage assignment was limited without reference cell-type information. Triwise comparisons across disease states demonstrated that deep learning–derived features accounted for the strongest disease-associated signal, with several features enriched in tumor samples. Feature distributions were largely concordant between paired nostrils, supporting aggregation of intra-patient data. These results demonstrated disease-associated morpholomic differences but underscored the need for a lineage-informed framework to enable cell-type–anchored interpretation.

### Differential morpholomic features distinguish health, chronic inflammation, and cancer

Deep learning–derived morpholomic features were used to generate an integrated low-dimensional embedding of all nasal swab–derived cells, visualized by UMAP (Figure 1b). Clustering within this shared morpholomic space revealed groups of cells with distinct size, shape, and texture characteristics (Figures 1b–1f). Several clusters composed of smaller, circular cells were inferred to represent immune populations, whereas clusters containing larger cells with eccentric morphologies were hypothesized to represent epithelial populations. In a subset of cells, elongated morphologies with apical appendage-like structures were observed, consistent with features typically associated with ciliated epithelial cells. However, while gross cellular classes could be inferred from morphologic characteristics, precise immune or epithelial subtypes could not be reliably assigned based on morphology alone.

To assess whether disease-associated morpholomic differences were detectable at a global level, we performed triwise comparisons across healthy controls, CRS, and sinonasal carcinoma using z-score–normalized feature values aggregated across samples (Figure 1g). This analysis revealed that deep learning–derived features were the most differentially represented across disease states. Several features exhibited preferential enrichment in tumor samples, whereas others were more strongly associated with CRS or non-tumor conditions. Comparison of cluster-level distributions between matched left and right nostrils from the same individuals demonstrated largely concordant cellular compositions (Supplementary Figure 1), indicating minimal intra-patient spatial variability. Modest discrepancies were observed in a small number of samples with lower cellular yield from one nostril, likely reflecting effects of downsampling required for computational processing of this type of data. To maximize statistical power, data from paired nostrils were therefore combined for subsequent analyses.

Statistical testing performed on patient-level median feature values further identified multiple deep learning–derived features and select texture-based features as differentially represented between health, CRS, and carcinoma (Figure 1h). One deep learning feature (DL52) was consistently identified across both triwise and pairwise analyses and exhibited a higher-value subpopulation in tumor samples relative to CRS (Figure 1i). Mapping this feature back onto the morpholomic UMAP revealed localized regions of elevated DL52 signal (Figure 1j), suggesting association with specific cellular states rather than diffuse sample-level effects. Notably, some highly differential imaging features may have been more related to sample processing rather than intrinsic cellular identity. These limitations motivated construction of a lineage-resolved reference atlas to constrain interpretation to biologically grounded cellular state. These findings demonstrate that disease-associated morpholomic features are detectable in nasal swabs but highlight the need for a lineage-informed reference framework. We therefore next sought to construct a single-cell immune morpholomic atlas to enable cell-type–anchored interpretation of nasal swab morpholomics.

### Lineage-resolved immune morpholomics define a reference atlas for nasal cytology

To enable lineage-informed interpretation of nasal swab morpholomics, we next constructed the first single-cell immune morpholomic reference atlas using purified immune cell populations with known identities. Commercially available, purified immune cells spanning both myeloid and lymphoid lineages were fixed and independently processed through the REM-I platform under identical imaging conditions (Figure 2a). Seven immune cell types were selected based on their anticipated morphologic diversity, including differences in cell size, texture, and shape, providing a structured foundation for downstream atlas-based analyses.

**Figure 2:**
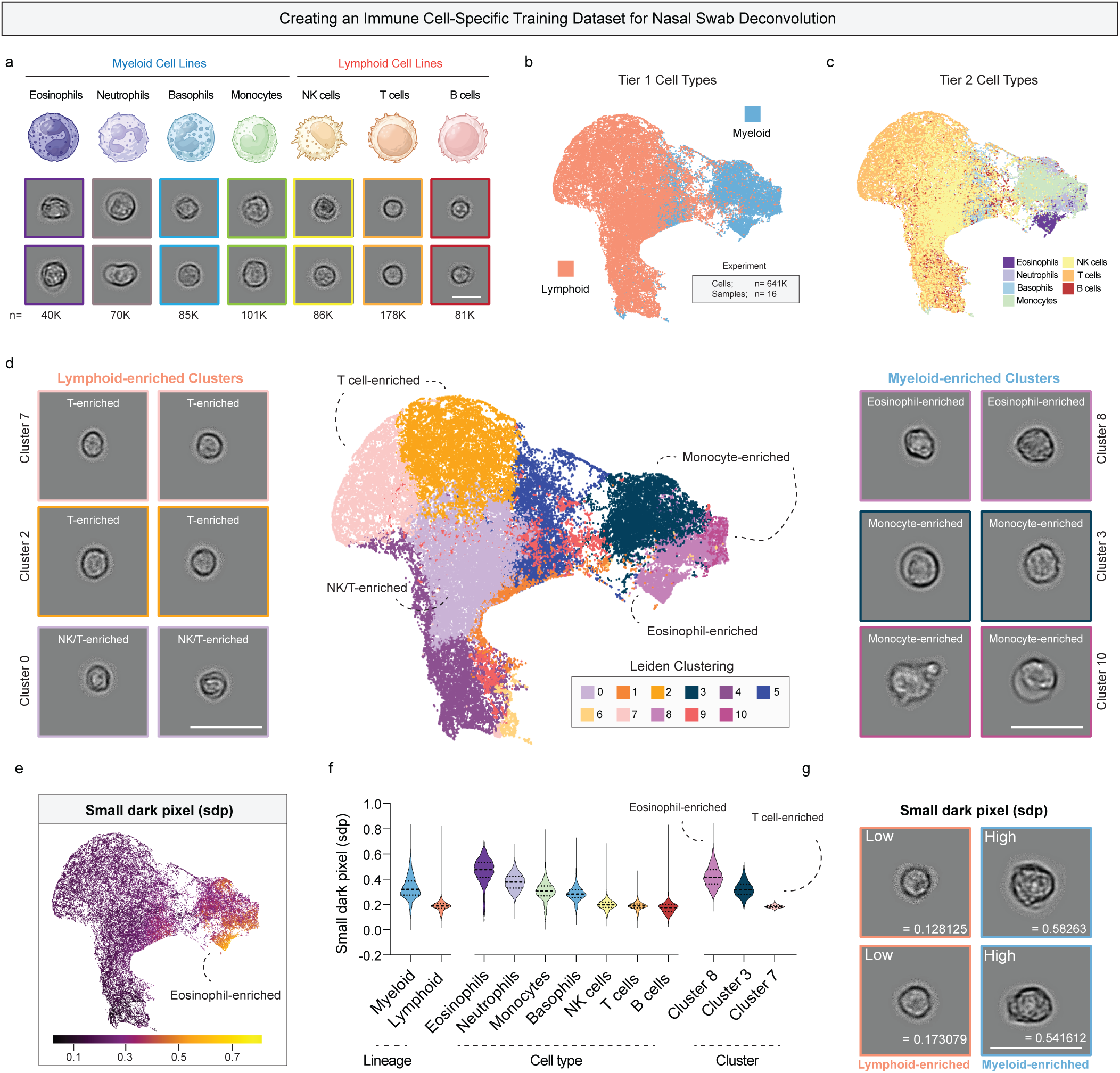
Construction of a lineage-resolved immune cell morpholomic atlas for nasal swab deconvolution. (a) Representative brightfield images of the seven purified immune cell populations used to generate the reference morpholomic atlas, spanning myeloid and lymphoid lineages. Number of cell images obtained for each cell type is listed below images. Scale bar represents 10µm. (b) UMAP embedding of all immune cells colored by Tier 1 lineage classification (myeloid vs lymphoid). (c) UMAP of immune cells colored by Tier 2 immune cell identity. (d) Unsupervised Leiden clustering of immune cells with representative images from select clusters; cluster identities were annotated post hoc using purified cell overlays. Scale bar represents 20µm. (e) UMAP colored by small dark pixel intensity (SDP), a texture-based morpholomic feature. (f) Distribution of SDP values stratified by immune lineage, cell type, and select clusters, demonstrating higher variance and enrichment within myeloid populations. (g) Representative cell images illustrating low and high SDP values, corresponding to granule-poor and granule-rich cellular morphologies, respectively. Scale bar represents 20µm. Illustrations from a) were created with BioRender.com

Integration of all immune cell feature embeddings revealed robust Leiden clustering driven by immune lineage (Figure 2b; Supplementary Figure 2a). Across the full dataset, 95.0% of lymphoid cells localized to 7 of 11 clusters, whereas 88.5% of myeloid cells were concentrated within the remaining 4 clusters, indicating strong morpholomic separation between major immune compartments. At finer resolution, several clusters were dominated by specific immune cell types. For example, cluster 7 was enriched for T cells (89.7%), while cluster 10 was primarily composed of monocytes (89.1%) (Figure 2c; Supplementary Figure 2b,c). Eosinophils exhibited particularly strong clustering behavior, with 89.3% localizing to a single cluster (cluster 8), which also contained most neutrophils (67.3%), reflecting shared granulocytic morphologies.

Inspection of representative cell images from lineage-enriched clusters revealed consistent intra-cluster morphologic features, particularly among lymphoid populations, which exhibited relatively uniform size and shape characteristics (Figure 2d). Consistent with this observation, no cluster was uniquely enriched for natural killer (NK) cells, highlighting partial morphologic overlap among lymphoid subsets. To quantitatively assess morphologic distinctiveness, we computed Jensen–Shannon divergence (JSD) scores comparing each immune cell type to the remainder of the immune population (Supplementary Table 1), as well as comparing individual clusters to all others (Supplementary Table 2). Myeloid populations, particularly neutrophils and eosinophils, demonstrated high divergence scores (>0.6), indicating strongly distinct morpholomic feature distributions. Among lymphoid cells, T cells exhibited the highest divergence score (0.51), consistent with moderate but incomplete separability. Together, these results demonstrate that immune lineage and subtype are reflected in morpholomic feature space, with greater discriminatory power for myeloid than lymphoid populations, and establish a quantitative, lineage-resolved immune morpholomic atlas suitable for anchoring interpretation of complex nasal swab datasets.

Small dark pixel intensity (SDP), defined as the sum of pixel intensities in regions identified as being small dark structures (8 pixels or smaller), was the top differential feature identified using Jensen-Shannon Divergence in both eosinophils and NK cells, and was in the top five features for six out of the seven immune populations. We therefore further explored this feature and found that while it was uniformly low in cells of lymphoid origin, SDP varied greatly in myeloid cell populations (Figure 2e-g) making it useful for assigning cell types to clusters. Based on the definition of SDP and its presence in myeloid cells, we hypothesized that this feature related to the presence of granules in the cell and higher SDP values may correlate to higher granulocyte activity. This lineage-grounded immune morpholomic atlas establishes a transferable reference for cytologic analysis across diverse biofluids and mucosal tissues that contain heterogeneous immune and epithelial cell populations, including inhalation interface biofluids such as saliva and bronchoalveolar lavage as an example.

### Integrated immune–nasal morpholomic atlas enables higher-resolution cell typing of nasal swabs

As the reference immune cell morpholomic atlas demonstrated lineage-specific feature structure, we next sought to leverage this information to improve cell-type annotation within heterogeneous nasal swab samples. We therefore integrated the purified immune cell atlas with the nasal swab morpholomic dataset, generating a unified atlas spanning both healthy and disease-associated cellular states (Figure 3a,b). Leiden clustering was performed on the integrated dataset, and clusters containing fewer than 100 cells were excluded to ensure analytical robustness (Figure 3c). Overlaying reference immune cell embeddings onto the integrated atlas revealed clear spatial segregation between immune-enriched and epithelial-enriched regions, with two distinct immune-dominated compartments emerging within morpholomic space. Two clusters (clusters 2 and 8) contained cells that bridged immune- and epithelial-enriched regions (Supplementary Figure 3a). Visual inspection of representative cell images from these clusters demonstrated distinct differences in size and shape, supporting the presence of multiple underlying populations; epithelial-enriched clusters (2 and 8) and procedure-associated contaminant clusters (Supplementary Figure 3b,c). Using this 12-cluster framework, we examined representative images, key morpholomic features (log-transformed area, eccentricity, small dark pixel intensity, and mean pixel intensity), and the spatial overlap with reference immune populations to annotate cluster identities (Figure 3d–h; Supplementary Figure 3d,e).

**Figure 3:**
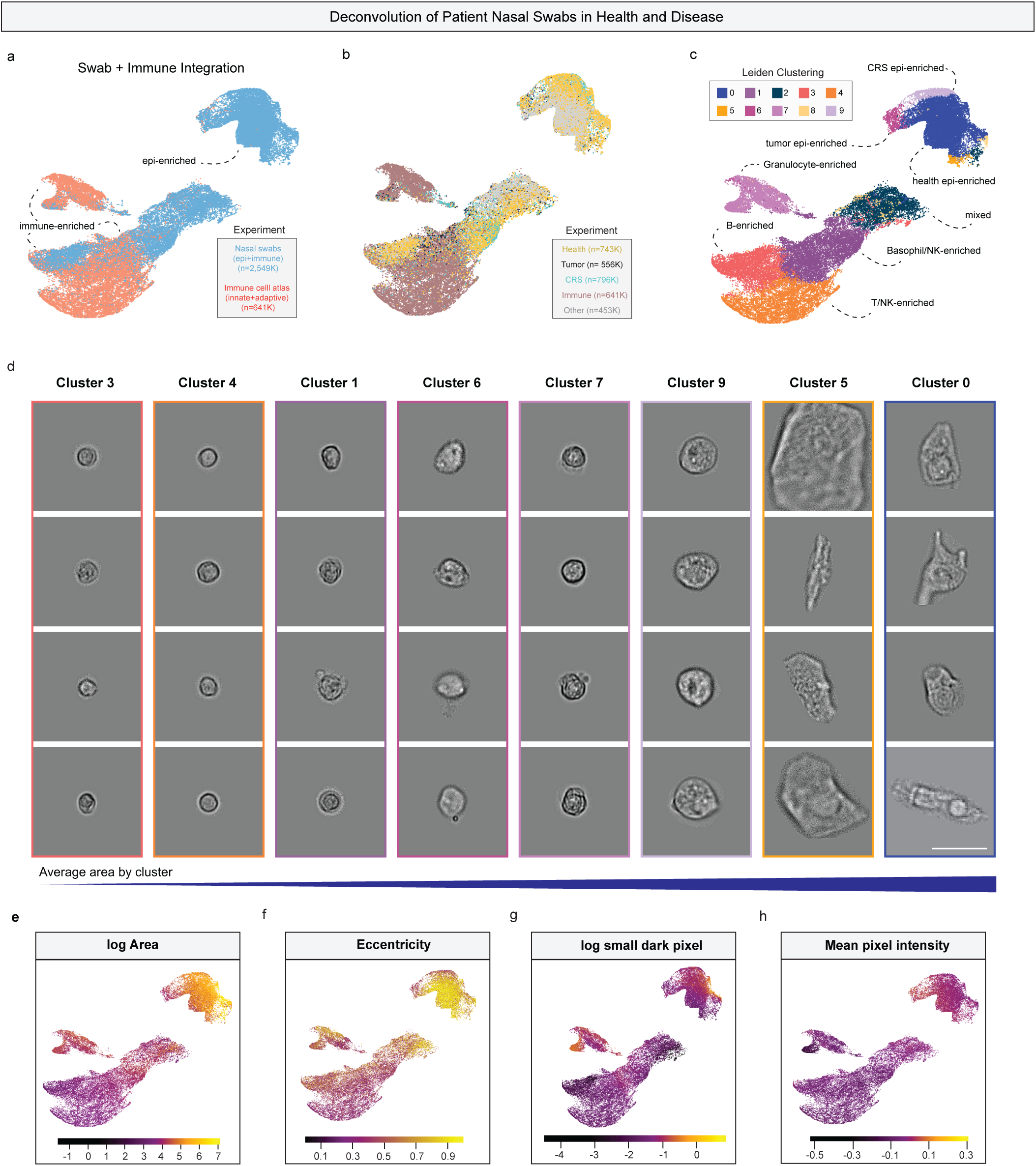
Integrated reference immune and nasal swab morpholomic atlas enables deconvolution of nasal cell populations. (a) UMAP of the integrated morpholomic atlas combining purified reference immune cells and nasal swab–derived cells, colored by dataset of origin. (b) UMAP colored by clinical disease state, demonstrating overlap of healthy, chronic rhinosinusitis (CRS), and tumor samples within shared cellular states. (c) UMAP colored by unsupervised Leiden clustering performed on the integrated morpholomic space. (d) Representative brightfield images from each Leiden cluster, ordered by increasing average cell area, illustrating morphologic distinctions between immune-enriched and epithelial-enriched populations. Scale bar represents 20µm. (e–h) UMAPs colored by key morpholomic features: (e) log-transformed cell area, (f) eccentricity, (g) log-transformed small dark pixel intensity, and (h) mean pixel intensity. Feature gradients align with immune–epithelial separation and support cluster-level annotation.

Clusters 1, 3, 4, and 7 were annotated as basophil/NK-enriched, B cell–enriched, T/NK cell–enriched, and myeloid-enriched, respectively. Overlay of purified immune cells revealed that more than 75% of monocytes, neutrophils, and eosinophils localized to the myeloid-enriched cluster (cluster 7), precluding confident assignment of a single granulocytic subtype. However, the lower-left region of this cluster exhibited elevated small dark pixel intensity values consistent with eosinophil-associated morphology, suggesting that higher-resolution clustering may further resolve granulocyte substructure (Figure 3g). As other clusters did not demonstrate comparable internal heterogeneity, and to avoid over-partitioning, we chose to retained the 12-cluster solution for downstream analyses. Clusters 0, 2, 5, 6, 8, and 9 were clearly separated from immune-enriched regions and were defined by larger cell size and increased eccentricity, consistent with epithelial morphology. We therefore classified these clusters as epithelial-enriched and analyzed them independently for disease-associated epithelial features.

The procedure-associated contaminant clusters localized proximal to immune-enriched regions but did not overlap with reference immune cell embeddings. Cells within these clusters exhibited sizes comparable to immune cells but eccentricity values closer to epithelial cells, raising the possibility of non-nasal origin. Examination of clinical metadata revealed that these clusters were enriched in samples where nasal swabbing caused minor mucosal disruption and visible bleeding (Supplementary Figure 4). Based on these observations, we concluded that these clusters most likely represent blood-associated contaminants introduced during sampling, rather than bona fide nasal epithelial or immune populations. These clusters were therefore excluded from all subsequent analyses, and immune- and epithelial-enriched populations were carried forward separately for disease-specific morpholomic interrogation.

### Myeloid cell enrichment and activation distinguish tumor-associated immune states

Quantification of immune cell composition on a per-sample basis revealed a significant enrichment of myeloid-like cells in tumor samples compared with healthy controls (Mann Whitney test; Figure 4a). Although several chronic rhinosinusitis (CRS) samples also exhibited elevated myeloid-like fractions, this increase did not reach statistical significance, reflecting substantial inter-patient heterogeneity that may be related to differences in inflammatory burden or disease activity. Within the myeloid-enriched cluster, small dark pixel intensity (SDP) was significantly elevated in tumor-associated cells relative to both healthy and CRS samples (Figure 4b). Given prior analyses linking SDP to granulocyte-associated morphologic features, these findings indicate that tumor samples are characterized not only by increased myeloid cell abundance but also by changes to granulocytic activity, consistent with shifts to their innate immune state.

**Figure 4:**
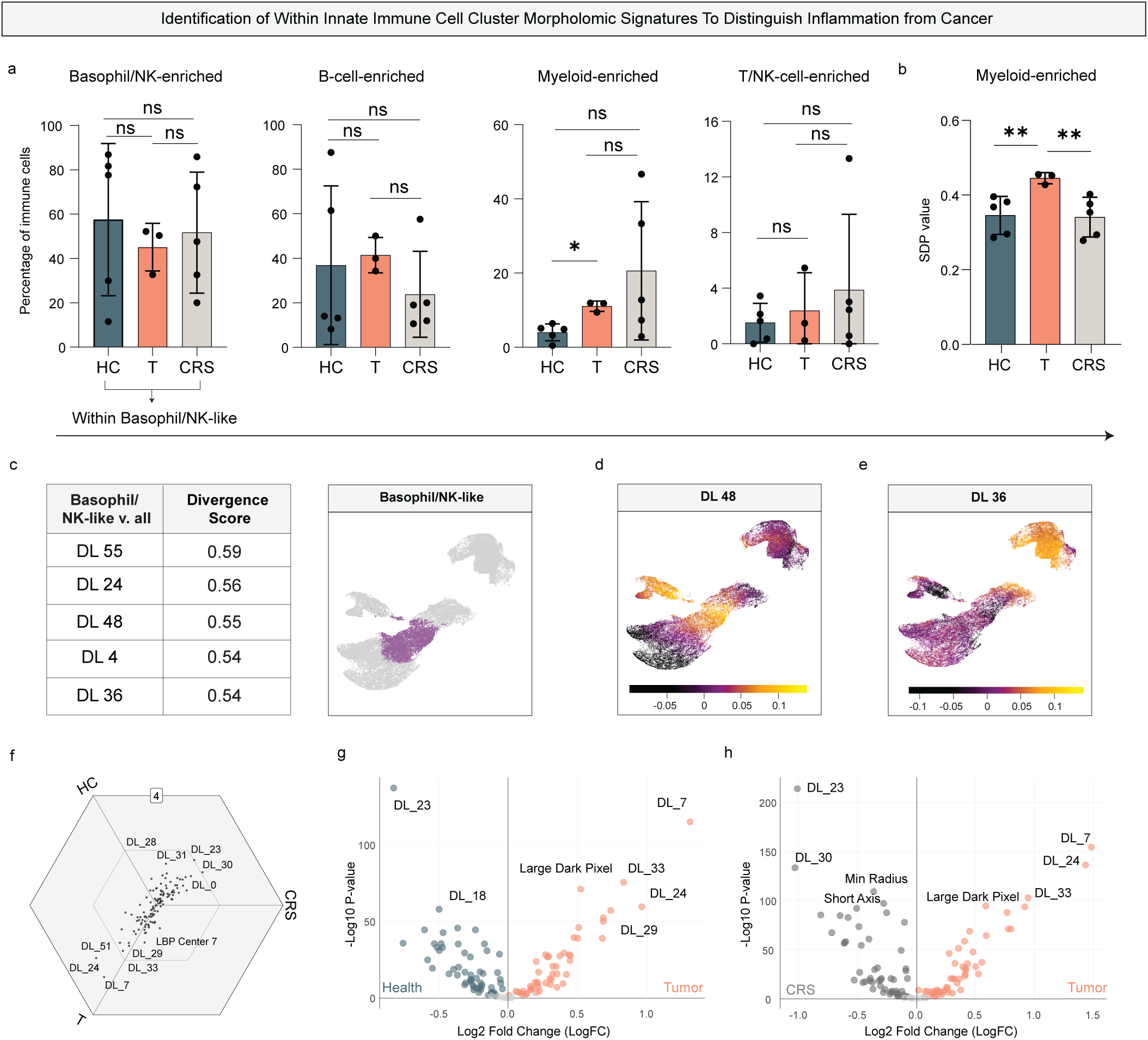
Within-lineage immune morpholomic signatures distinguish malignancy from chronic inflammation. (a) Proportion of immune-enriched Leiden clusters across healthy controls (HC, n=5), sinonasal carcinoma (T, n=3), and chronic rhinosinusitis (CRS, n=5). Statistical significance assessed using Mann Whitney test (*p < 0.05). Most immune populations show no significant differences (ns, non-significant) in relative abundance, indicating that cell frequency alone does not distinguish disease states. (b) Small dark pixel intensity (SDP) within myeloid-enriched cells across disease groups, demonstrating significantly elevated SDP in tumor-associated myeloid cells (Welch’s t-test, **p < 0.01). (c) Top five morpholomic features ranked by Jensen–Shannon divergence for basophil/NK-like cells relative to all other cells, highlighting deep learning–derived features as dominant discriminators. (d–e) UMAPs of nasal swab cells colored by representative deep learning features (DL48 and DL36), illustrating spatially localized, tumor-associated morpholomic states within basophil/NK-like populations. (f) Triwise comparison of sample-level mean z-score–normalized features for basophil/NK-like cells across HC, CRS, and tumor samples, showing tumor-associated feature enrichment. (g–h) Volcano plots of basophil/NK-like cells comparing tumor versus health (g) and tumor versus CRS (h), demonstrating tumor-associated deep learning and texture-based feature signatures that are absent in benign inflammatory disease.

The basophil/NK-enriched cluster represented the largest immune population across healthy, CRS, and tumor samples. Jensen–Shannon divergence analysis demonstrated that deep learning–derived features accounted for the majority of discriminative signal within this population relative to other cells in morpholomic space (Figure 4c; Supplementary Table 3). Several features, including elevated deep-learning feature DL48 and reduced DL36, were shared between basophil/NK-enriched cells and the myeloid-enriched cluster, likely reflecting shared morphologic characteristics of basophils with other associated granulocytic populations. Despite this partial overlap, basophil/NK-enriched cells remained spatially distinct from myeloid-enriched cells on the UMAP, enabling focused downstream analysis.

To evaluate disease-associated morpholomic shifts within basophil/NK-like cells, we performed both triwise feature analysis (Figure 4f) and differential feature testing using limma (Figure 4g,h). Across both analyses, DL–derived features emerged as the dominant disease-associated signals, with elevated DL7, DL24, and DL33 uniquely characterizing basophil/NK-like cells from tumor samples. In contrast, basophil/NK-like cells from CRS and healthy samples lacked distinct disease-specific features and shared multiple morpholomic signatures. Collectively, these results demonstrate that basophil/NK-like cells acquire tumor-associated morpholomic phenotypes that are not observed in benign inflammatory conditions, highlighting the potential of immune morpholomic profiling to distinguish malignant from chronic inflammatory sinonasal disease.

### Tumor-associated epithelial cells exhibit disease-specific morpholomic features

Given that sinonasal carcinomas arise from epithelial tissues, we next asked whether epithelial morpholomic features could independently distinguish malignant from inflammatory disease states. Analysis of epithelial-enriched clusters (clusters 0, 2, 5, 6, 8, and 9) revealed no statistically significant differences in their relative abundance across healthy, CRS, and tumor samples (Figure 5a). This lack of proportional change likely reflects a combination of limited cohort size and heterogeneity in epithelial composition across disease states. In the absence of consistent cluster-level enrichment, we focused subsequent analyses on the aggregate epithelial cell population to assess disease-associated morpholomic alterations independent of cell frequency.

**Figure 5.**
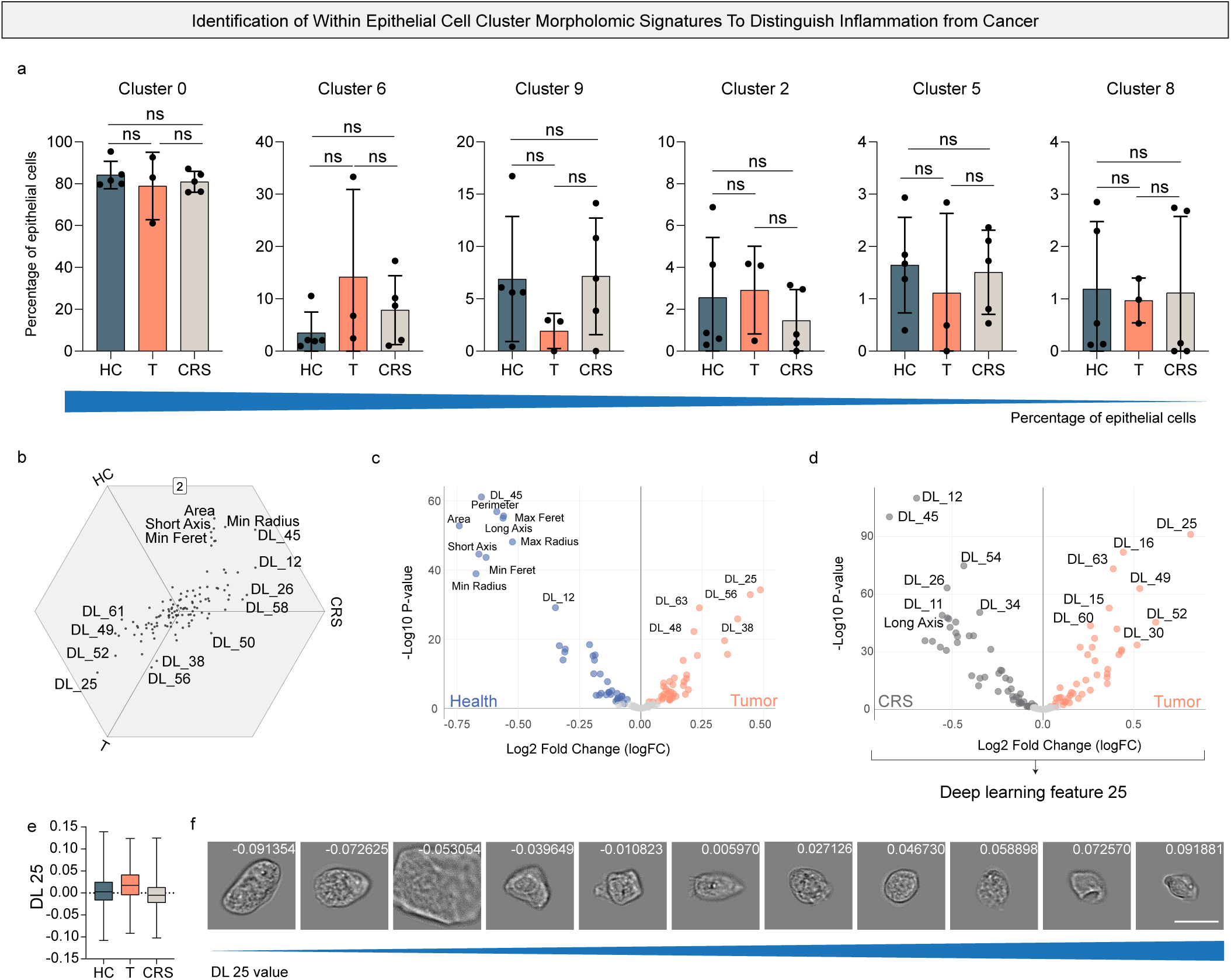
Morpholomic signatures of epithelial cells distinguish sinonasal carcinoma from chronic inflammation. (a) Proportion of epithelial-enriched cell clusters across healthy controls (HC, n=5), sinonasal carcinoma (T, n=3), and chronic rhinosinusitis (CRS, n=5). No significant differences in epithelial cluster abundance were observed across disease states using a Mann Whitney test (ns, non-significant). (b) Triwise comparison of epithelial morpholomic features between HC, CRS, and tumor samples using sample-level mean z-score normalization. c–d) Volcano plots of epithelial morpholomic features (z-score normalized), highlighting differential features between (c) tumor versus health and (d) tumor versus CRS samples. (e) Box plot of cells DL 25 value across healthy, tumor and CRS samples. (f) Select representative images of cells across a range of DL 25 values. Exact feature values are shown. Scale bar represents 20µm.

Triwise feature analysis and differential feature testing were therefore performed across all epithelial cells, comparing tumor samples with healthy controls and CRS (Figure 5b–d). Across both analytical approaches, tumor-associated epithelial cells were consistently characterized by reduced cell size, a finding that aligns with established histopathologic features of malignant epithelial transformation. Size reduction remained a dominant discriminative feature when comparing tumor epithelial cells to both healthy and CRS-associated epithelium, indicating that this morphologic shift is not attributable solely to chronic inflammation.

In addition to size-based differences, several deep learning–derived features were uniquely enriched in tumor epithelial cells. DL25 consistently appeared as higher in tumor samples vs healthy and CRS samples (Figure 5 b-e). Representative images of cells across DL25 values show changed in the size and texture, suggesting these are defining factors for the deep learning feature (Figure 5f). These observations were confirmed using histograms of select features across DL25 values (Supplemental Figure 5). Notably, elevated DL52, previously identified in bulk morpholomic analyses, was reproducibly associated with tumor epithelial cells in both triwise comparisons and volcano plot–based differential testing. These results demonstrate that malignant transformation within the nasal epithelium is accompanied by distinct, high-dimensional morpholomic remodeling that can be detected directly from minimally invasive nasal swabs. These findings indicate that epithelial morpholomic features, particularly reduced cell size and tumor-associated deep learning signatures, provide complementary diagnostic information to immune-based morpholomic states. Using morpholomic profiling of epithelial cell types with the most proportionally rich immune cell population enables multivariable characterization of sinonasal disease, supporting the utility of single-cell morpholomics for distinguishing malignancy from chronic inflammatory conditions in the upper airway.

### Disease-associated morpholomic feature signatures distinguish malignancy

For nasal swab morpholomic analysis to support clinical translation, disease-associated feature signatures must reliably distinguish malignant lesions from benign inflammatory masses without reliance on endoscopy, biopsy, or other invasive procedures. Although no statistically significant differences were observed in the proportion of basophil/NK-like cells across disease states, earlier analyses suggested that tumor-associated basophil/NK-like cells exhibit altered morpholomic states, consistent with changes in cellular activity rather than abundance. We therefore assessed whether disease-associated differences could be detected at the feature level on a per-sample basis. Median feature values were calculated for each sample, and Welch’s t-tests were performed to compare feature distributions across disease groups. Within basophil/NK-like cells, deep learning–derived and texture-based features emerged as the most statistically significant discriminators of tumor samples relative to both healthy controls and CRS (Figure 6a–e). Consistent with these findings, qualitative differences in cellular morphology were observed between tumor-associated and non-tumor basophil/NK-like cells (Figure 6c,e). Substantial variability in some median feature values was observed across CRS samples, likely reflecting heterogeneity in inflammatory burden and disease activity (Figure 6b). While several features distinguished either health versus tumor or CRS versus tumor, values of the morphometric feature LBP Center 10 were consistently higher in healthy and CRS samples than in tumors, identifying this feature as a potential negative marker for malignancy (Figure 6d).

**Figure 6:**
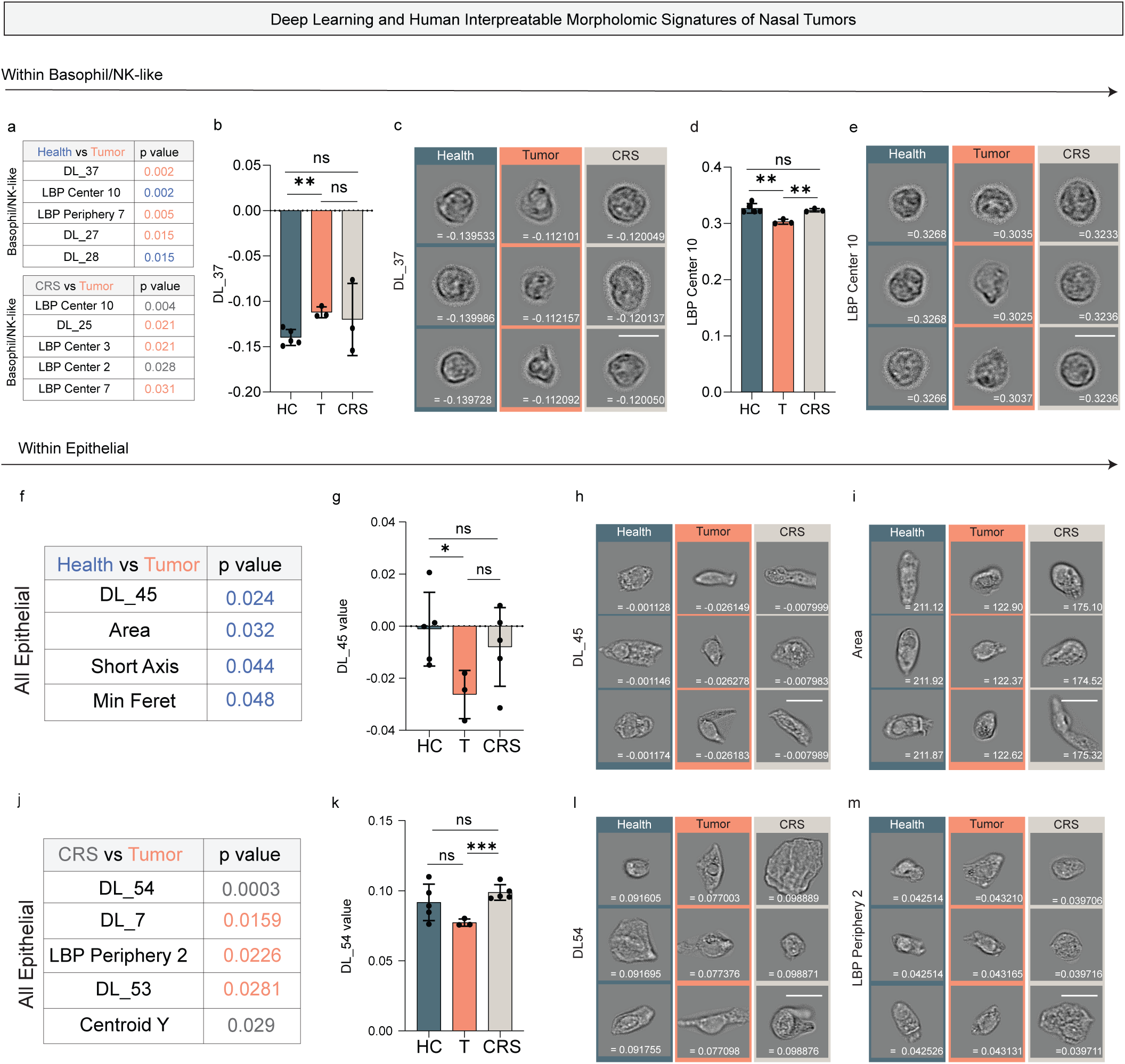
Deep learning and human-interpretable morpholomic signatures distinguish sinonasal carcinoma from chronic inflammation. (a) Top differentially expressed morpholomic features in basophil/NK-like cells identified by Welch’s t-test comparing health (HC, n=5) versus tumor (T, n=3) and CRS (n=3) versus tumor samples. Feature colors indicate higher relative abundance in the indicated condition. (b) Median values of deep learning feature DL37 in basophil/NK-like cells across health (HC, n=5), sinonasal carcinoma (T, n=3), and chronic rhinosinusitis (CRS, n=3). Statistical significance determined using Welch’s t-test. **p < 0.01, ns=non-significant. (c) Representative brightfield images of basophil/NK-like cells with near-median DL37 values from each disease group; exact feature values are shown. Scale bar represents 10µm. (d) Median values of LBP Center 10 in basophil/NK-like cells across HC (n=5), T (n=3), and CRS (n=3) samples. Statistical significance determined using Welch’s t-test. **p < 0.01, ns=non-significant. (e) Representative images of basophil/NK-like cells with near-median LBP Center 10 values; exact feature values are shown. Scale bar represents 10µm. (f) Top differentially expressed epithelial morpholomic features identified by Welch’s t-test when comparing healthy versus tumor samples. Feature colors indicate higher relative abundance in the indicated condition. (g) Median values of epithelial deep learning feature DL45 across HC (n=5), T (n=3), and CRS (n=3) samples. Statistical significance determined using Welch’s t-test. *p < 0.05, ns=non-significant. (h) Representative epithelial cells with near-median DL45 values across disease states; exact feature values are shown. Scale bar represents 20µm. (i) Representative epithelial cells with near-median area values across disease states; exact feature values are shown. Scale bar represents 20µm. (j) Top differentially expressed epithelial morpholomic features identified by Welch’s t-test when comparing CRS versus tumor samples. Feature colors indicate higher relative abundance in the indicated condition. (k) Median values of epithelial deep learning feature DL54 across HC (n=5), T (n=3), and CRS (n=3) samples. Statistical significance determined using Welch’s t-test. ***p < 0.001, ns=non-significant. (l) Representative epithelial cells with near-median DL54 values; exact feature values are shown. Scale bar represents 20µm. (m) Representative epithelial cells with near-median LBP Periphery 2 values; exact feature values are shown. Scale bar represents 20µm.

To build a more robust disease signature, we repeated this analysis across the aggregate epithelial cell population. This analysis confirmed a statistically significant reduction in median epithelial cell size in tumor samples compared with healthy controls and identified a deep learning–derived feature (DL45) that was significantly enriched in tumor-associated epithelial cells (Figure 6f–i). Although triwise analysis suggested that epithelial cells from healthy and CRS samples share similar size distributions, Welch’s t-tests did not identify epithelial size as a highly discriminative feature between these two benign conditions. In contrast, deep learning–derived features and the texture-based feature LBP Periphery 2 were among the most statistically significant discriminators between health and CRS, indicating that subtle epithelial morpholomic remodeling accompanies chronic inflammation even in the absence of malignancy. These results demonstrate that statistically significant, disease-associated morpholomic feature signatures can be detected at the sample level from nasal swabs. By integrating immune and epithelial features, morpholomic analysis enables discrimination of sinonasal carcinoma from benign inflammatory disease, supporting its potential role as a non-invasive potential point-of-care diagnostic test in the future for stratifying patients between inflammatory conditions and malignant conditions in clinically ambiguous sinonasal pathology.

## DISCUSSION

Early detection of sinonasal malignancies remains a major clinical challenge. These tumors are rare, frequently present with nonspecific symptoms, and overlap clinically with more common chronic inflammatory conditions such as chronic rhinosinusitis (CRS) with nasal polyposis, resulting in delayed diagnosis and poorer outcomes. Screening strategies that are rapid, cost effective, minimally invasive, and deployable in primary care settings could identify patients who require advanced imaging, biopsy, and subspecialty evaluation. Given that the nasal cavity is both a commonly afflicted site of sinonasal tumors and easily accessible in clinical practice^20^, nasal swabs represent an attractive opportunity for cellular-level diagnostic assessment.

In this study, we demonstrate that deep learning–enabled single-cell morpholomics can extract clinically relevant immune and epithelial signatures from routine nasal swabs without staining or complex sample preparation. Using the REM-I platform, we performed high-throughput, label-free brightfield imaging of cells obtained from healthy individuals, patients with CRS, and individuals with biopsy-confirmed sinonasal carcinoma. Despite the absence of molecular labels, deep learning–derived morpholomic features consistently emerged as the most discriminative signals across analyses, suggesting that high-dimensional morphology alone contains sufficient information to resolve disease-associated cellular states.

A central challenge in interpreting nasal swab morpholomics was the absence of established ground-truth references for cell-type assignment. To address this, we first constructed a lineage-resolved immune morpholomic atlas using purified immune populations with known identities. This reference atlas revealed that immune lineage, particularly myeloid versus lymphoid identity, is strongly encoded in morpholomic feature space, with myeloid populations exhibiting the greatest intra- and interpopulation separability. This finding is consistent with traditional cytology methods including H+E staining, which uses nuclear morphology and cytoplasmic granules to distinguish granulocytes and lymphoid cells^21^. However, as our method does not use cellular staining, small dark pixel intensity (SDP), a feature associated with dense intracellular structures, emerged as a dominant discriminator of granulocytic populations and was hypothesized to reflect granule-associated morphology. Overall, developing this immune atlas provided a necessary foundation for interpreting heterogeneous nasal swab data and enabled higher-confidence annotation of immune-enriched populations in disease samples.

Integration of the immune reference atlas with nasal swab morpholomics revealed specific disease-associated remodeling of immune cell states. Tumor samples exhibited both an increased proportion of myeloid-like cells and elevated SDP within these cells, suggesting heightened granulocytic activity in the tumor-associated immune milieu. Basophil/NK-enriched populations also demonstrated tumor-associated deep learning feature signatures that were not present in CRS or healthy samples, indicating that immune perturbations in malignancy involves qualitative cellular changes beyond simple shifts in abundance. This idea is supported by previous studies which have shown that the actin cytoskeleton of NK cells facilitates functions including infiltration, granule secretion and cell binding, and will be remodeled as part of a tumor response^22–24^. Therefore, cellular architecture can be altered in tumor-associated inflammation and immune morpholomics may detect those changes and capture functional states relevant to malignancy.

Epithelial morpholomics provided a complementary disease signal. Although epithelial cluster proportions did not differ significantly across disease states, tumor-associated epithelial cells consistently exhibited reduced cell size and distinct deep learning feature profiles compared with both CRS and healthy controls. Importantly, these findings are compatible with what brightfield morpholomics can robustly measure, such as cell size, shape, and texture, without relying on nuclear segmentation or staining-dependent features such as nucleus-to-cytoplasm ratio. While classical histopathologic assessment of sinonasal carcinomas emphasizes nuclear atypia, keratinization, and architectural disarray, many of these features are not directly observable in unstained single-cell brightfield images. Instead, our results suggest that malignant epithelial remodeling, consistent with previously seen cancer cytoskeleton responses to immune invasion, manifests as reproducible, high-dimensional morpholomic shifts that are detectable even in the absence of molecular or histologic labels^25^.

From a diagnostic perspective, the combined immune and epithelial signatures identified here support the feasibility of morpholomic cytology as a screening tool rather than a replacement for all histopathology. At the sample level, statistically significant differences in deep learning and texture-based features distinguished tumor samples from benign inflammatory conditions. Features such as reduced epithelial cell size, altered deep learning embeddings, and immune-associated texture metrics (e.g., LBP features) may form the basis of a future composite diagnostic signature capable of flagging malignant lesions for expedited workup and referral.

### Study limitations

This study has several limitations. First, the cohort size is modest, reflecting the rarity of sinonasal malignancies, and larger, multi-institutional datasets will be required to validate and generalize these findings. Second, while deep learning features are highly discriminative, their biological interpretability remains limited, underscoring the need for continued integration with orthogonal assays such as molecular profiling or targeted staining in future studies. Third, brightfield morpholomics cannot directly assess nuclear features traditionally used in histopathologic diagnosis, and conclusions are therefore appropriately constrained to measurable cellular morphology and population-level patterns.

Future work will focus on expanding cohort size, refining predictive feature sets, and training supervised classifiers capable of producing probabilistic malignancy scores with defined confidence intervals. Advancements in the real-time sorting capabilities of the REM-I further open the possibility of isolating morpholomically defined populations for downstream genomic or transcriptomic analysis, allowing for more insights into the tumor biology. Together, these advances position single-cell morpholomic cytology as a promising, scalable approach for early, point-of-care diagnosis of sinonasal disease and potentially other mucosal tissues characterized by mixed immune and epithelial populations.

## METHODS

### Immune atlas generation

To generate a morpholomic atlas of healthy immune cells, sorted, cryopreserved populations of immune cells were purchased from the following vendors: Stanford Blood Center (B cells, CD4/CD8 T cells (labelled T cells in this data), NK cells, monocytes), and CGT Preclinical (CD14+ monocytes, Basophils, Neutrophils, Eosinophils). Two unique patient samples were obtained for each immune population. Samples were thawed according to vendor recommendations, resuspended in Deepcell sample buffer to a final concentration between 1 x 10^6^ to 3 x 10^6^ cells/mL, and imaged on the REM-I platform (Deepcell) using REM-I imaging chips. Between about 5,000 and 55,000 images were obtained for each sample, at sample throughputs between about 10 cells/s and 100 cells/s.

### Clinical cohort

Nasal brush samples were collected using a cytobrush (Medscand Cytobrush Plus) from patients with either suspected or confirmed nasal carcinoma, chronic rhinosinusitis (CRS), or patients with no known nasal diseases (health) at the University of North Carolina Hospitals. Suspected carcinomas were confirmed via biopsy and patient clinical and demographic information can be found in Table 1. All patients provided informed consent as part of an approved IRB (# UNC-IRB-17-2677, PI: Kimple). The study included individuals 18 years or older who underwent nasal endoscopy for inflammatory or non-inflammatory pathology. To be included in the malignancy cohort, patients needed a biopsy-confirmed sinonasal malignancy diagnosed either before or after sample collection. Patients were included in the inflammatory cohort if they met clinical criteria for CRS and demonstrated endoscopic evidence of sinonasal inflammation, as confirmed by a board-certified otolaryngologist. Control patients were those undergoing endoscopic nasal procedures for conditions unrelated to sinonasal malignancy or inflammation (e.g., deviated nasal septum, endoscopic endonasal pituitary surgery.) Informed consent was obtained from patients in the perioperative area of a tertiary care surgical hospital prior to their scheduled procedure.

### Sample collection

Sample collection was performed after patients were under general anesthesia and positioned safely for their planned surgical procedures. Nasal epithelial samples were collected separately from each nostril of the patient using a sterile cytobrush. The cytobrush was gently inserted approximately 2-3 cm posterolaterally, directed along the inferior turbinate, and rotated 3-5 times to collect epithelial cells. Each cytobrush was then carefully withdrawn and immediately placed into a pre-labeled individual 50ml conical tube containing 7 mL of PBS. Conicals were placed on ice until samples could be transported to the lab for further processing.

### Sample processing

PBS from the tube was collected via a pipette and used to thoroughly wash the brush, removing the sample from the brush. 1 mL of 16% paraformaldehyde (PFA) was added to each tube to reach a final concentration of 2% PFA. Sample was mixed, transferred to a 15 mL conical tube and allowed to fix for 20 minutes at room temperature on a rocker set to a low, gentle rocking speed. After 20 minutes, the sample was diluted with 6 ml of PBS and centrifuged at 800g for 5 minutes at room temperature. The supernatant was removed from the sample post centrifugation, and the sample was washed by resuspending the pellet in 6 mL of PBS and centrifuging again. After two washes had occurred, the final pellet was resuspended in 1 ml of PBS and stored in a labelled cryovial tube at 4 C.

### Nasal sample imaging on the REM-I platform

Nasal samples filtered with a 70um filter to remove larger cells which could clog the imaging chip. Each solution was centrifuged and resuspended in 250µl- 350µl of Deepcell sample buffer. Samples were imaged on the REM-I at a rate of about 5 cells/s to 50 cells/s. For each sample, about 10,000 to 300,000 images were collected on the REM-I, which was based on the total cell number in each sample

### Data filtering and UMAP generation

Data was collected and compiled in the Axon Data Suite (Deepcell) to create a nasal swab only atlas, an immune reference atlas, and an integrated immune/nasal swab atlas. For each high-resolution cell image captured on the system, the REM-I platform extracts 115 morphological features (64 Deep-learning features and 51 Computer Vision Human Interpretable features) based on the Human Foundation Model (HFM)^26,27^. All data sets were downsampled proportionally to 100,000 cells total for processing. UMAPs based on the deep-learning morphological features were generated using 0.5 Leiden resolution, while the UMAP hyperparameters of minimum distance and spread were set to 0.1 and 1 respectively. Samples were then filtered to remove cellular debris and Deepcell focusing beads, resulting in a “nasal swab only” atlas of 81,308 cells (16 Leiden clusters), a “final immune reference” atlas of 62,436 cells (11 Leiden clusters), and an “integrated” atlas of 52,989 cells (13 Leiden clusters). All morphological features for each cell with corresponding metadata were exported as a .tsv file for subsetting and analysis. Clusters with less than 100 cells were removed from downstream analysis. In the integrated atlas, clusters with large proportions of known immune cells from the reference atlas were assigned as immune dominant clusters and separated out for further analysis in figure 3. 2 cell clusters were found to be split across different locations in the UMAP and therefore were further subclustered into two separate clusters. One of each subclusters was found to be the dominant cell population in samples from swabs that produced blood, suggesting these cells were a contaminating population and were therefore removed from immune and epithelial cell analysis. The remaining cell clusters were composed of suspected epithelial cells and were separated out for further analysis in figure 4.

### Differential feature analysis and triwise analysis on normalized cell populations

To perform differential feature analysis and generate volcano and triwise plots, each morphological feature had to be z-score normalized by applying the formula Z= (X-mu)/ sigma where X= individual feature value (per cell), mu= mean value of feature across all cells, sigma= standard deviation of feature across all cells. Cells were subset based on assigned clusters and disease state (Chronic Rhinosinusitis (CRS), Tumor (T) and Healthy control (HC)) for analysis. Differential feature analysis was performed using the limma package in R studio (version 4.3.3) which outputted log2 fold change (logFC) of the z-scores and raw p-values^28^. Multiple testing correction was applied using the Benjamini-Hochberg procedure. Volcano plots were then generated using these values, with reported features colored by directionality and the top proteins (ranked by nominal P-value) labeled.

For comparison of all three disease states at once, the triwise package was used in R studio. Mean Z score values of each feature across each condition were imputed to get barycentric coordinates which were then plotted in 2D. Each feature is represented by a single dot, with the direction of the dot indicating largest z-scores.

### Statistical analysis and graph generation

All statistical analysis, heatmaps, violin plots, box plots, and bar charts were generated using GraphPad Prism V10.4.1. Mann Whitney U-test was used for testing cell proportion differences of clusters between disease types. For bar charts examining differences in features across disease type, the median feature value for each sample was used. If multiple samples were run from the same patient, e.g.: samples collected from both nostrils, the median values were then averaged to get a final value for each patient. Comparison of cell type and features across disease type was performed using Welch’s t-test. * indicate a p-value <0.05 and ** indicates a p-value <0.01.

**Supplementary Table 1:** Jensen-Shannon Divergence results comparing each purified immune cell type to all other immune cell populations.

**Supplementary Table 2:** Jensen-Shannon Divergence results comparing each cluster in reference immune atlas to all other cell populations.

**Supplementary Table 3:** Jensen-Shannon Divergence results comparing each cluster in integrated immune and nasal atlas to all other cell populations.

**Supplementary figure 1:** Intra-patient consistency of nasal swab morpholomic composition.

Heatmap showing the percentage of cells assigned to each Leiden cluster from independently processed left and right nostril swabs for individual CRS patients. Total numbers of cells analyzed per swab on the Axon platform are shown below each column. Only swabs without reported contamination (e.g., blood or fungal material) and from patients with bilateral clinical diagnoses were included. Overall, cluster distributions were largely concordant between nostrils within the same patient, indicating limited intra-patient spatial variability.

**Supplementary figure 2:** Lineage and cell-type composition across immune cell Leiden clusters.

a. Proportion of lymphoid and myeloid cells within lymphoid-dominated clusters (Leiden clusters 0, 1, 2, 4, 6, 7, and 9) and myeloid-dominated clusters (Leiden clusters 3, 5, 8, and 10).
b. Relative contribution of individual immune cell types (eosinophils, neutrophils, basophils, monocytes, B cells, T cells, and NK cells) within lymphoid-dominated and myeloid-dominated clusters.
c. Distribution of each immune cell type across all Leiden clusters, shown as the percentage of cells per cluster.

**Supplementary figure 3:** Characterization of intermediate clusters in the integrated immune and nasal atlas.

a. Leiden clustering of the integrated reference immune and nasal swab atlas highlighting the emergence of intermediate clusters following subdivision of clusters 2 and 8.
b. Representative brightfield images of cells from both epithelial-enriched and intermediate clusters 2 and 8 demonstrating distinct differences in cell size and morphology.
c. Box plot showing cell area distributions across the split clusters, illustrating that intermediate (inter.) clusters 2 and 8 are significantly smaller than their epithelial (epi.)-enriched counterparts.
d. Heatmap showing the percentage contribution of each sample type (healthy control, CRS, tumor, tumor-adjacent, and purified immune populations) across all clusters, grouped into immune-like, intermediate, and epithelial-like categories.
e. Heatmap of select median morpholomic feature values for each cluster, including mean pixel intensity, eccentricity, log small dark pixel intensity, LBP texture features, maximum radius, and representative deep learning features.

**Supplementary figure 4: Comparison of cellular distributions between contaminated and uncontaminated nasal swabs.**

Density plots of cells projected onto the integrated UMAP for six nasal swabs collected from three patients, each contributing one visibly bloody (contaminated) and one non-bloody (uncontaminated) swab. Contaminated swabs show localized enrichment of cells within intermediate regions of morpholomic space, whereas uncontaminated swabs display minimal occupancy of these regions. These patterns support the interpretation that specific intermediate clusters arise from blood-derived contamination rather than nasal-resident cell populations.

**Supplementary figure 5: Analysis of feature changes across DL 25 values.** Histogram showing changes in select features across bins of DL 25 values. a) Median LBP Periphery 8 values, b) Median Area, and c) Median of cells’ mean pixel intensity values.

## Resource availability

### Lead contact

Further information and resource requests should be directed to the lead contact, Kevin Matthew Byrd.

### Materials availability

This study did not generate new unique reagents.

### Data and code availability

All original code has been deposited at https://github.com/Loci-lab

## Data Availability

All data produced in the present study are available upon reasonable request to the authors

## Acknowledgements

We thank all patients who generously provided samples for this study. We also thank the team at the University of North Carolina Marsico Lung Institute Respiratory TRACTS Core, particularly Mandy Bush and Griselda Portillo, for their assistance with fixation, storage, and shipment of nasal swab samples. We are grateful to Tony Gonzalez for technical support with processing nasal swabs on the REM-I platform. We additionally thank the Deepcell team for their technical guidance and support throughout this study, and the ADA Science and Research Institute for institutional support that enabled this work. Biorender.com was used to generate figure 1a and figure 2a.

The authors acknowledge funding support from the American Dental Association Science and Research Institute (Startup funds), the Chan Zuckerberg Initiative (2021-237918), and Virginia Commonwealth University (Startup funds from Philips Institute for Oral Health Research and Massey Comprehensive Cancer Center, and Massey Cancer Center Harrison Scholars Award).

## Author contributions

Conceptualization: BTR, AJ, JM, AK, KMB

Data curation: BTR, AJ, KS, JM

Formal analysis: BTR, AJ, KS

Funding acquisition: AK, KMB

Investigation: BTR, AJ, TW, KS, MB, JM, QTE

Methodology: BTR, AJ, TW, KS, MB

Project administration: AJ, AK, KMB

Resources: AJ, KS, MB, JM, AK, KMB

Software: BTR, KS

Supervision: AK, KMB

Validation: BTR, AJ, JM

Visualization: BTR, AJ

Writing – original draft: BTR, AJ, JM, AK, KMB

Writing – review & editing: BTR, AJ, TW, KS, MB, JM, QTE, AK, KMB

