## Supplementary figures and images for "A Deep Learning–Enabled Single-Cell Morpholomic Atlas of Nasal Swabs Distinguishes Chronic Inflammation from Sinonasal Malignancy"

### Supplemental Figure 5

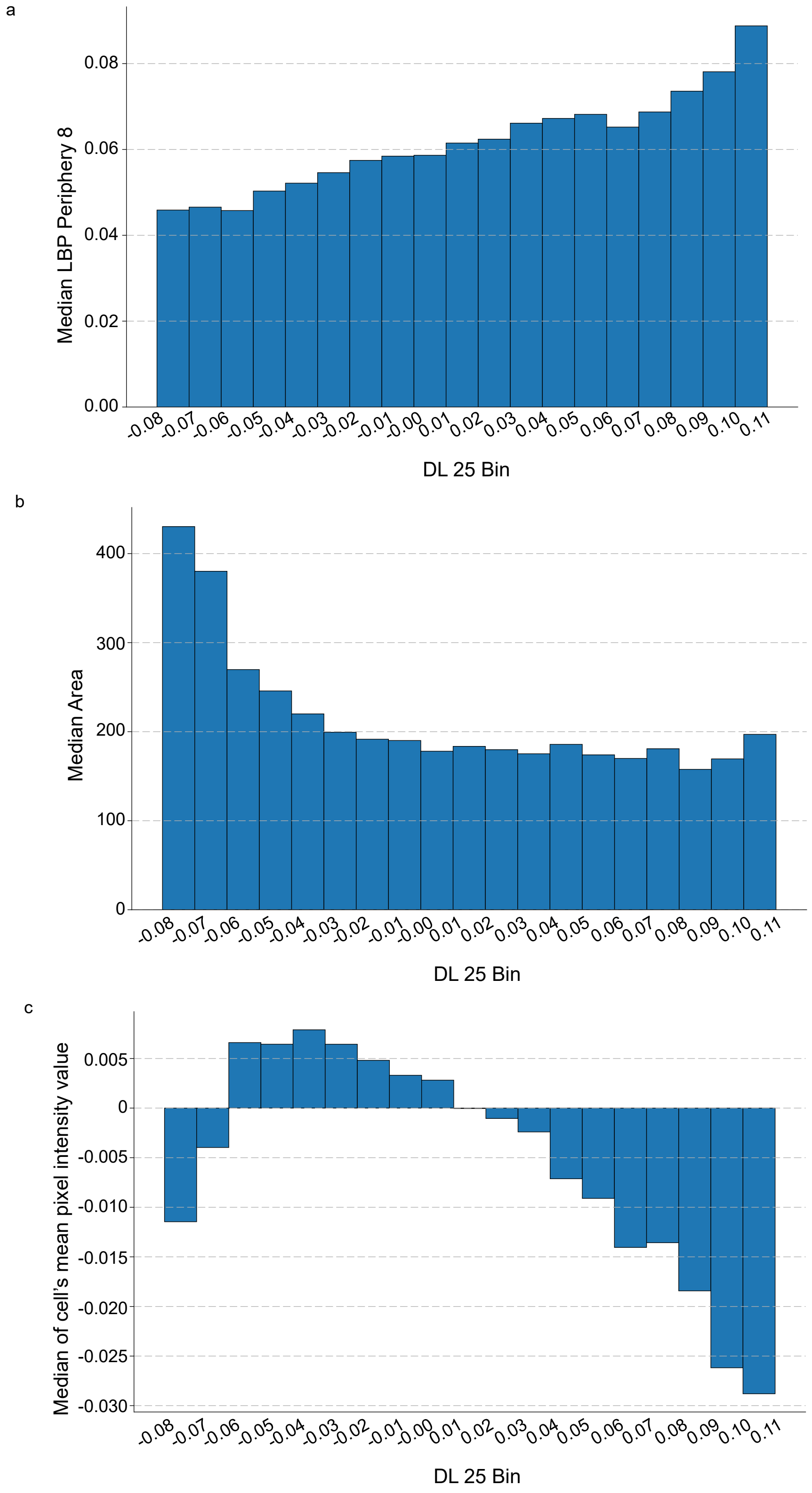

### Supplementary Figure 1

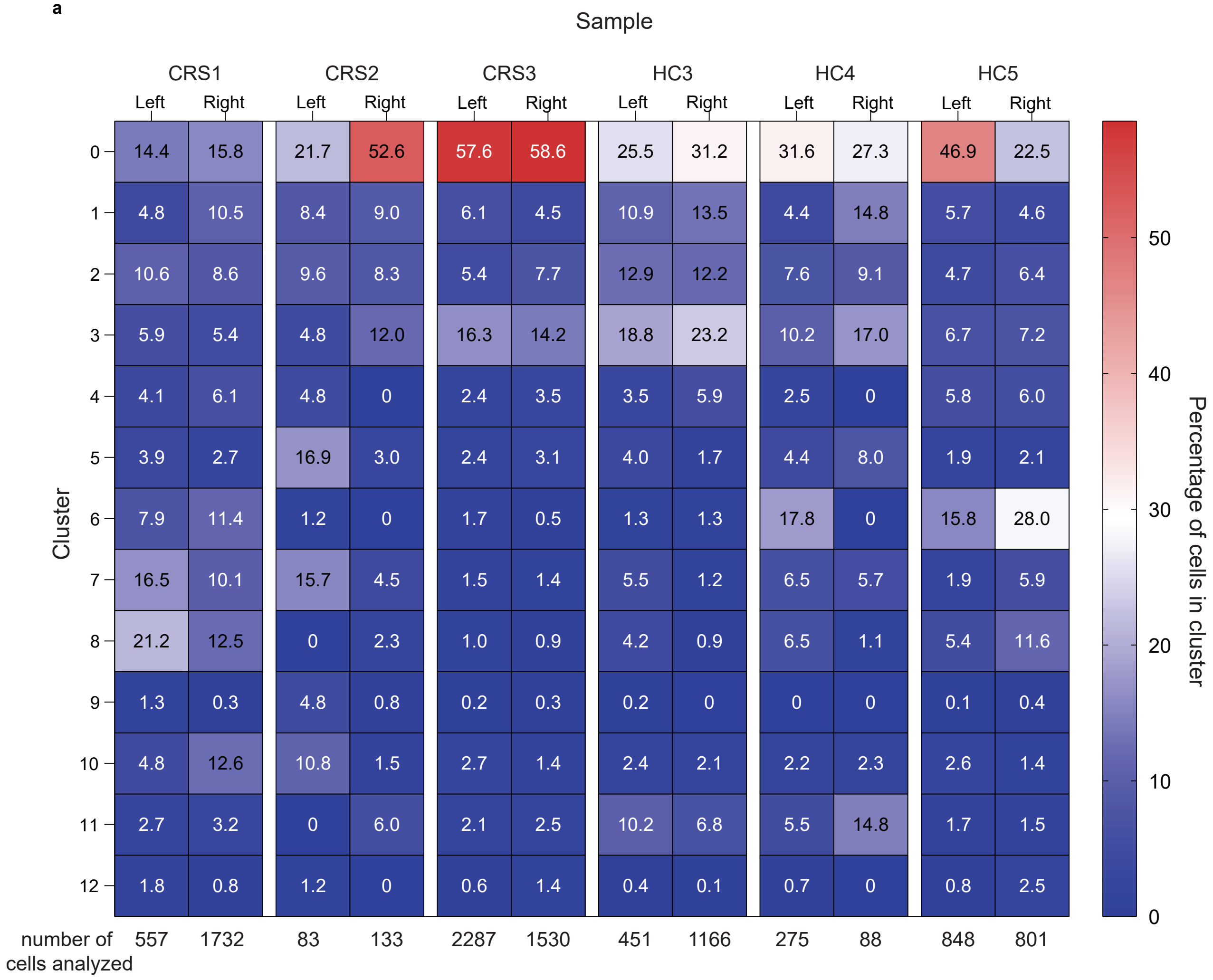

### Supplementary Figure 2

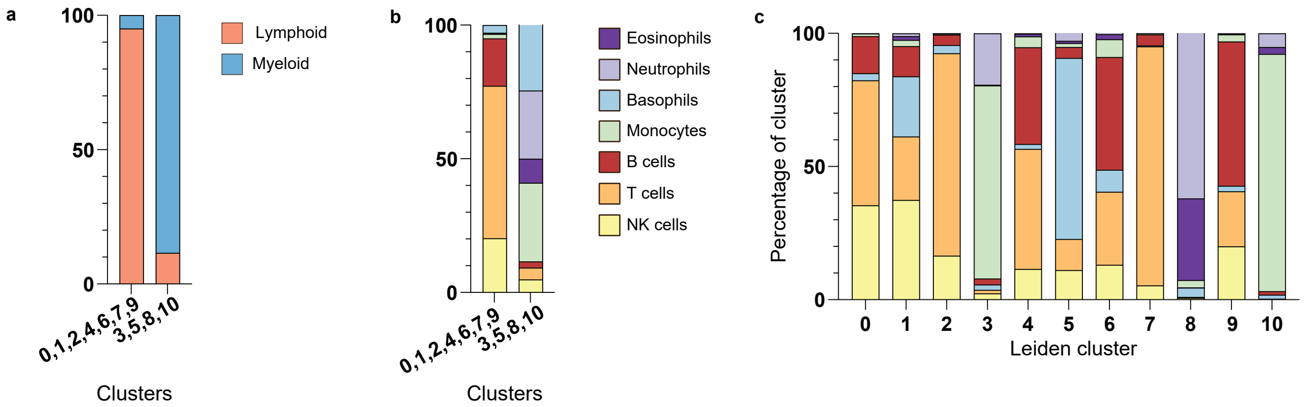

### Supplementary Figure 4

Supplemental figure 4s

**Density plots of bloody vs non-blood nostril from same patient**

a

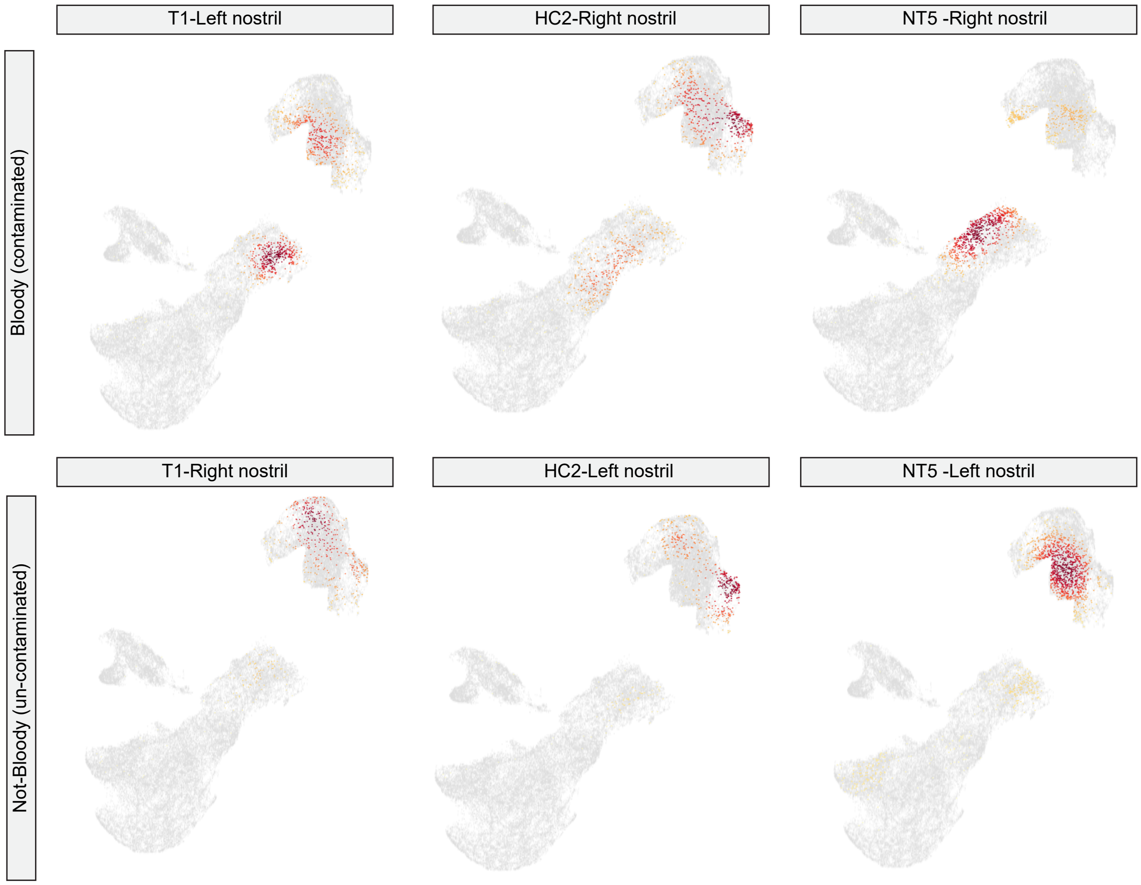
