## Supplementary Figure 3 for "A Deep Learning–Enabled Single-Cell Morpholomic Atlas of Nasal Swabs Distinguishes Chronic Inflammation from Sinonasal Malignancy"

Figure 3s

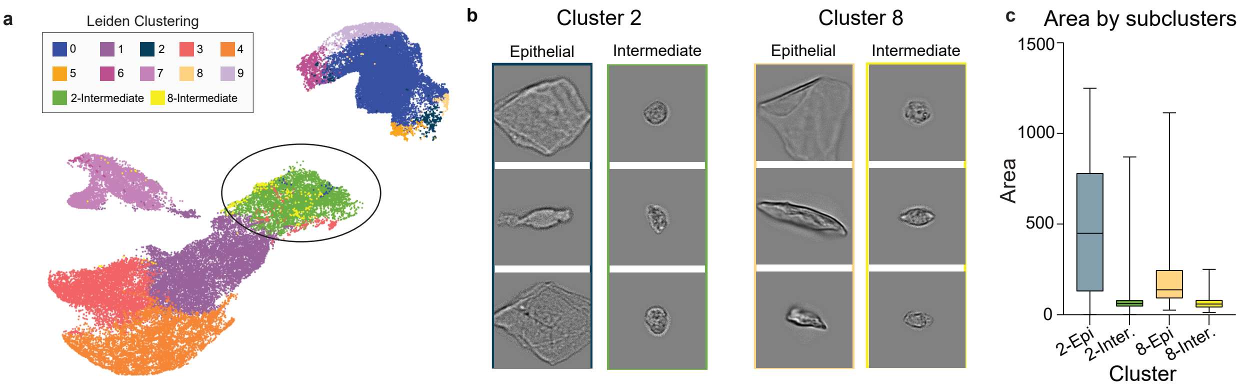


d

| | Immune-like | | | | Intermediate | | Epithelial-like | | | | | |
|---|---|---|---|---|---|---|---|---|---|---|---|---|
| | 7 | 3 | 1 | 4 | 2 | 8 | 6 | 5 | 8 | 9 | 0 | 2 |
| HC | 0.81 | 14.60 | 13.23 | 0.45 | 15.50 | 0.64 | 2.09 | 0.92 | 0.51 | 3.81 | 46.31 | 1.12 |
| CRS | 1.84 | 4.64 | 25.01 | 0.41 | 10.56 | 0.38 | 1.86 | 0.52 | 0.35 | 5.55 | 48.34 | 0.53 |
| T | 2.87 | 11.62 | 13.34 | 0.38 | 29.79 | 1.91 | 3.20 | 0.81 | 0.29 | 1.05 | 33.43 | 1.29 |
| Tum Adj | 0.30 | 1.50 | 1.28 | 0 | 9.70 | 0.15 | 2.33 | 1.88 | 1.73 | 10.23 | 66.47 | 4.44 |
| Monocytes | 75.53 | 3.71 | 5.40 | 2.02 | 5.68 | 0.09 | 5.26 | 0.05 | 0 | 0.19 | 1.64 | 0.42 |
| Eosinophlis | 88.33 | 1.21 | 6.28 | 1.69 | 0.95 | 0.05 | 0.37 | 0 | 0.16 | 0.05 | 0.90 | 0 |
| Neutrophils | 91.60 | 0.27 | 5.39 | 1.44 | 0.45 | 0.04 | 0.49 | 0 | 0 | 0.04 | 0.27 | 0 |
| B cells | 1.96 | 55.20 | 22.28 | 14.29 | 4.34 | 0.53 | 0.98 | 0 | 0.03 | 0.03 | 0.36 | 0 |
| Basophils | 3.95 | 2.55 | 67.16 | 15.42 | 8.88 | 0.32 | 1.26 | 0 | 0 | 0.04 | 0.40 | 0.04 |
| NK | 0.90 | 18.70 | 46.02 | 32.60 | 1.31 | 0.20 | 0.15 | 0 | 0.09 | 0 | 0.03 | 0 |
| T cells | 0.35 | 22.10 | 13.62 | 63.16 | 0.41 | 0.10 | 0.11 | 0 | 0 | 0 | 0.14 | 0 |

(Color scale: 0, 20, 40, 60, 80)

e

| | | | | | | | | | | | | |
|---|---|---|---|---|---|---|---|---|---|---|---|---|
| Mean pixel intensity | -0.073 | -0.025 | -0.069 | -0.086 | -0.025 | -0.028 | 0.033 | 0.007 | -0.032 | 0.004 | 0.001 | -0.001 |
| Eccentricity | 0.54 | 0.50 | 0.46 | 0.36 | 0.61 | 0.75 | 0.51 | 0.87 | 0.88 | 0.56 | 0.83 | 0.71 |
| log small dark pixel | -0.946 | -1.929 | -1.409 | -1.641 | -1.773 | -1.602 | -1.382 | -1.931 | -1.087 | -1.078 | -1.126 | -1.731 |
| LBP Periphery 3 | 0.022 | 0.028 | 0.022 | 0 | 0.061 | 0.048 | 0.050 | 0.067 | 0.041 | 0.034 | 0.059 | 0.054 |
| Max radius | 5.758 | 4.011 | 4.823 | 3.866 | 5.520 | 5.887 | 5.476 | 11.760 | 11.056 | 5.887 | 12.094 | 17.376 |
| DL 35 | -0.0719 | 0.0368 | -0.0295 | 0.0684 | 0.0004 | -0.0201 | 0.0521 | 0.0563 | -0.0136 | 0.1160 | 0.0777 | 0.0205 |

(Color scales: Mean pixel intensity 0, -0.05; Eccentricity 0.8, 0.6, 0.4; log small dark pixel -1.2, -1.6; LBP Periphery 3 0.04, 0; Max radius 15, 5; DL 35 0.1, 0)
