## Supplementary Table 1 for "A Deep Learning–Enabled Single-Cell Morpholomic Atlas of Nasal Swabs Distinguishes Chronic Inflammation from Sinonasal Malignancy"

| Features | | Divergence score | Features | | Divergence score |
| --- | --- | --- | --- | --- | --- |
| B cells | |  | T cells | |  |
| DL 21 | | 0.41 | DL 5 | | 0.51 |
| DL 9 | | 0.38 | DL 43 | | 0.50 |
| Small Dark Pixels | | 0.38 | DL 49 | | 0.49 |
| Large Dark Pixels | | 0.37 | Small Dark Pixels | | 0.49 |
| DL 08/ DL 58/  Pixel Int. 25^th^ perc./ Stdev | | 0.35 | DL 36 | | 0.48 |
| NK | |  | Monocytes | |  |
| Small Dark Pixels | | 0.34 | DL 49 | | 0.57 |
| DL 5 | | 0.34 | DL 23 | | 0.55 |
| Small Bright Pixels | | 0.33 | DL 3 | | 0.52 |
| Large Bright Pixels | | 0.33 | DL 34 | | 0.52 |
| DL 14 | | 0.31 | DL 64 | | 0.52 |
| Eosinophils | |  | Basophils | |  |
| Small Dark Pixels | | 0.62 | Small Dark Pixels | | 0.42 |
| Large Dark Pixels | | 0.57 | DL 5 | | 0.42 |
| Std Pixel Intensity | | 0.57 | DL 48 | | 0.41 |
| Image moment 1 | | 0.56 | DL 58 | | 0.41 |
| Small Bright Pixels/ DL 27 | | 0.56 | DL 16/ DL 49 | | 0.40 |
| Neutrophils | |  |  |  | |
| DL 5 | | 0.63 |  |  | |
| DL 23 | | 0.61 |  |  | |
| Small Dark Pixels | | 0.60 |  |  | |
| Small Bright Pixels | | 0.60 |  |  | |
| DL 52/ DL 1 | | 0.59 |  |  | |
