## Supplementary Table 2 for "A Deep Learning–Enabled Single-Cell Morpholomic Atlas of Nasal Swabs Distinguishes Chronic Inflammation from Sinonasal Malignancy"

| Features | Divergence score | Features | Divergence score |
| --- | --- | --- | --- |
| Leiden 0 |  | Leiden1 |  |
| DL 33 | 0.53 | DL 36 | 0.49 |
| DL 23 | 0.53 | DL 49 | 0.47 |
| DL 19 | 0.53 | DL 62 | 0.46 |
| DL 9 | 0.52 | DL 55 | 0.44 |
| DL 49 | 0.52 | DL 16 / DL34 | 0.43 |
| Leiden 2 |  | Leiden 3 |  |
| DL 61 | 0.67 | DL 49 | 0.70 |
| DL 44 | 0.64 | DL 23 | 0.67 |
| DL 31 | 0.60 | DL 31 | 0.66 |
| DL 62 | 0.58 | DL 36 | 0.66 |
| DL 39 /DL 3 | 0.58 | DL 64 | 0.65 |
| Leiden 4 |  | Leiden 5 |  |
| DL 4 | 0.70 | DL 48 | 0.59 |
| DL 8 | 0.70 | DL 5 | 0.57 |
| DL 58 | 0.65 | DL 58 | 0.53 |
| DL 41 | 0.62 | DL 49 | 0.53 |
| DL 9 | 0.61 | Small Bright Pixels | 0.51 |
| Leiden 6 |  | Leiden 7 |  |
| DL 41 | 0.67 | DL 50 | 0.73 |
| DL 58 | 0.66 | DL 55 | 0.70 |
| DL 18 | 0.66 | DL 49 | 0.69 |
| DL 8 / DL 35 | 0.63 | DL 24 | 0.66 |
| Mean pixel intensity | 0.63 | DL 36/ DL62 | 0.65 |
| Leiden 8 |  | Leiden 9 |  |
| Small Bright pixel | 0.71 | DL 22 | 0.68 |
| DL 5 | 0.69 | Large Dark Pixels | 0.52 |
| Small Dark Pixel | 0.68 | Mean Pixel Intensity | 0.49 |
| DL 40 | 0.68 | DL 11 | 0.49 |
| DL 14 | 0.65 | DL 64 | 0.49 |
| Leiden 10 |  |  |  |
| DL 6 | 0.75 |  |  |
| DL 48 | 0.73 |  |  |
| DL 28 | 0.69 |  |  |
| DL 13 | 0.68 |  |  |
| Small Bright Pixel/ DL 12/ DL 14 | 0.66 |  |  |
