## Supplementary Table 3 for "A Deep Learning–Enabled Single-Cell Morpholomic Atlas of Nasal Swabs Distinguishes Chronic Inflammation from Sinonasal Malignancy"

| Features | | Divergence score | Features | | | | Divergence score |
| --- | --- | --- | --- | --- | --- | --- | --- |
| Leiden 0 | |  | Leiden1 | | | |  |
| DL 35 | | 0.73 | DL 55 | | | | 0.59 |
| Max radius | | 0.70 | DL 24 | | | | 0.56 |
| Max caliper dist. | | 0.70 | DL 48 | | | | 0.55 |
| Long ellipse axis | | 0.69 | DL 4 | | | | 0.54 |
| Perimeter | | 0.68 | DL 36 | | | | 0.54 |
| Leiden 2 | |  | Leiden 3 | | | |  |
| DL 13 | | 0.58 | DL 9 | | | | 0.66 |
| DL 31 | | 0.55 | DL 27 | | | | 0.59 |
| Large dark pixels | | 0.54 | DL 5 | | | | 0.58 |
| DL 35 | | 0.53 | Small dark pixels | | | | 0.55 |
| DL 12 | | 0.50 | DL 61/ Small bright pixel | | | | 0.54 |
| Leiden 4 | |  | Leiden 5 | | | |  |
| DL 61 | | 0.72 | DL 58 | | | | 0.70 |
| LBP Periphery 3 | | 0.64 | DL 35 | | | | 0.68 |
| DL 62 | | 0.62 | DL 42 | | | | 0.68 |
| Long ellipse axis | | 0.61 | DL 19 | | | | 0.68 |
| Perimeter | | 0.61 | Negative fraction/DL22/DL59 | | | | 0.67 |
| Leiden 6 | |  | Leiden 7 | | | |  |
| DL 28 | | 0.70 | DL 36 | | | | 0.68 |
| DL 6 | | 0.66 | DL 33 | | | | 0.64 |
| DL 60 | | 0.65 | DL 52 | | | | 0.64 |
| DL 7 | | 0.63 | DL 63 | | | | 0.63 |
| DL 61 | | 0.63 | DL 1/ DL5 | | | | 0.61 |
| Leiden 8 | |  | Leiden 9 | | | |  |
| DL 13 | | 0.59 | DL 6 | | | | 0.67 |
| DL 62 | | 0.53 | DL 12 | | | | 0.66 |
| DL 32 | | 0.51 | DL 20 | | | | 0.63 |
| DL 36 | | 0.51 | Min radius | | | | 0.63 |
| DL 25 | | 0.51 | Short ellipse axis | | | | 0.63 |
